# A Systematic Review: The Dimensions utilized in the Performance Evaluation of Healthcare- An Implication during the COVID-19 Pandemic

**DOI:** 10.1101/2021.06.26.21259568

**Authors:** Faten Amer, Sahar Hammoud, Haitham Khatatbeh, Szimonetta Lohner, Imre Boncz, Dóra Endrei

## Abstract

**Background:** The balanced scorecard (BSC) has been implemented to evaluate the performance of health care organizations (HCOs). BSC proved to be effective in improving financial performance and patient satisfaction.

**Aim:** This systematic review aims to identify key performance indicators (KPIs) and dimensions that are vital and most frequently used by health care managers in BSC implementations. Additionally, it attempts to analyze the resulting dimensions during the COVID-19 era.

**Methods:** This systematic review adheres to PRISMA guidelines. The PubMed, Embase, Cochrane, and Google Scholar databases and Google search engine were inspected to find all implementations of BSC at HCO. The risk of bias was assessed using the nonrandomized intervention studies (ROBINS-I) tool to evaluate the quality of observational and quasi-experimental studies and the Cochrane (RoB 2) tool for randomized controlled trials (RCTs).

**Results:** There were 33 eligible studies, of which we identified 36 BSC implementations. The categorization and regrouping of the 797 KPIs resulted in 46 subdimensions. The reassembly of these subdimensions resulted in 13 major dimensions: financial, efficiency and effectiveness, availability and quality of supplies and services, managerial tasks, health care workers (HCW) scientific development error-free and safety, time, HCW-centeredness, patient-centeredness, technology, and information systems, community care and reputation, HCO building, and communication. On the other hand, this review detected that BSC design modification to include external and managerial perspectives was necessary for many BSC implementations.

**Conclusion:** This review solves the KPI categorization dilemma. It also guides researchers and health care managers in choosing dimensions for future BSC implementations and performance evaluations in general. Consequently, dimension uniformity will improve the data sharing and comparability among studies. Additionally, despite the pandemic negatively influencing many dimensions, the researchers observed a lack of comprehensive HCO performance evaluations. In the same vein, although some resulting dimensions were assessed separately during the pandemic, other dimensions still lack investigation. Lastly, BSC dimensions may play an essential role in tackling the COVID-19 pandemic. However, further research is required to investigate the BSC implementation effect in mitigating the pandemic consequences on HCO.

## 1. Introduction

Evaluating the health care sector is quite challenging and complex. Unsatisfactory performance can result from long waiting times (WT), inefficiency, dissatisfactory patients, and health care workers’ (HCW) burnout [1, 2]. Coronavirus disease 2019 (COVID-19) imposed further burdens on the health care system worldwide due to the limited capacity of hospital beds and the increased psychological stress of HCWs during the COVID-19 pandemic [3, 4]. There is still a lack of information that would help health care managers and policymakers in the era of COVID-19 to improve the delivery of health care quality and to learn for the future [5]. Higher pandemic burdens, such as HCW burnout and stress, will rise when health care organizations (HCOs) lack plans and preparedness to strengthen their surge capacity and HCW resilience [6, 7].

Researchers employed different tools for the performance evaluation (PE) of HCO. The most utilized PE tools were the International Organization for Standardization (ISO standards), Malcolm Baldrige National Excellence Model (MBNQA), European Foundation for Quality Management (EFQM) Excellence Model, Singapore Quality Award (SQA), Six Sigma, Data Envelopment Analysis (DEA), Pabon Lasso Model, and Balanced Scorecard (BSC) [8–12].

The World Health Organization (WHO) initiated the Performance Assessment Tool for Quality Improvement in Hospitals (PATH) in 2003. It aimed to develop a framework for the assessment of hospital performance. The resulting dimensions from this project were clinical effectiveness, efficiency, HCW orientation, responsive governance, safety, and patient-centeredness. However, studies have shown that there are still some gaps in this model and issues concerning the dimensions investigated [13, 14]. Additionally, the Organization for Economic Co-operation and Development (OECD) launched the Health Care Quality Indicator (HCQI) project in 2006; it aimed to develop key performance indicators (KPIs) to compare quality in health care at the international level and achieve international benchmarking. This project concluded that health care must be safe, effective, patient-centered, timely, efficient, equitable, acceptable, and accessible [15, 16].

Most of the abovementioned managerial tools mainly focused on the KPIs related to quality, efficiency, productivity, and timeliness dimensions [8–12, 17]. Each of these dimensions is considered a dimension at the internal perspective of the BSC, which consists of four perspectives: the internal process, customer, innovation and growth perspectives, and financial perspectives [18]. Dimensions are described as collections of homogeneous or related KPIs. They are also referred to as diagnostic related groups (DRGs) [19], which have been proven to allow performance comparisons across hospitals and positively impact efficiency improvement [19].

The use of KPIs in the health care system before the pandemic has been beneficial for many reasons. First, the satisfaction rates of patients and HCWs were increased. Second, they lead to better efficiency, effectiveness, and financial performance and adapt to new technologies and ideas. Third, they lead to higher productivity and profitability [20–22]. In the pandemic, it is also crucial for HCO to track the performance of KPIs, which could draw faster attention to areas that require rapid responses and strengthening [6].

Most of the available PE models mainly focus on the internal perspective but lack coverage of the other dimensions or perspectives that are also important. BSC was considered different from the other managerial tools for two reasons. First, it offers a holistic approach to PE since it allows managers to highlight both financial and nonfinancial metrics. Second, the BSC is not only planning or a PE tool. It is also a strategic managerial tool that assigns KPIs compatible with the HCO strategy [23, 24]. However, other PE tools, such as total quality management (TQM), lack these comprehensive properties [25]. The first generation of the BSC, unveiled by Kaplan and Norton in 1992, involved four perspectives: the financial, customer, internal process, and innovation and growth perspectives, steered by the organizational vision and strategy [18]. See Figure (1).

**Figure 1:**
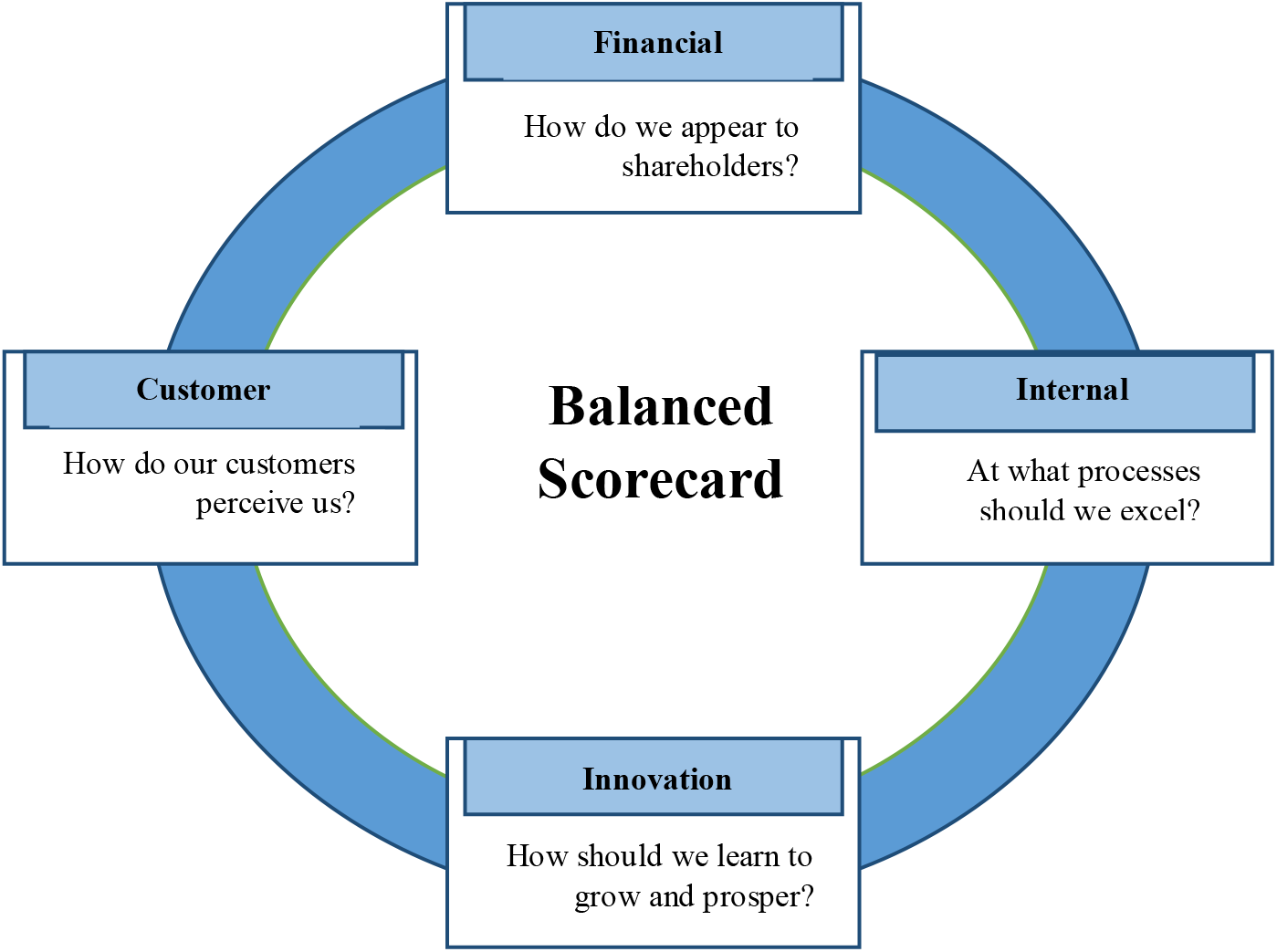
Balanced Scorecard Perspectives [18].

Later, the second generation of BSCs was developed to include strategic maps, in which cause-effect cascades between perspectives or KPIs were inspected [23]. In the third generation of BSCs, a destination statement was incorporated, which evokes where the organization plans to go within a time horizon and the action plans to achieve each targeted objective [24]. In health care, Duke Children’s Hospital in the United States of America (USA) was the first to implement the BSC in 1997. Figure (2) represents the strategic map of Duke University’s health system. As a result, the hospital converted 11 million American dollar losses into four million profits after four years of implementation [25]. Since then, BSC has gained increasing attention, and many HCO in high-income countries and low- and middle-income countries have strategically utilized BSC to develop their organizations [28–32].

**Figure 2:**
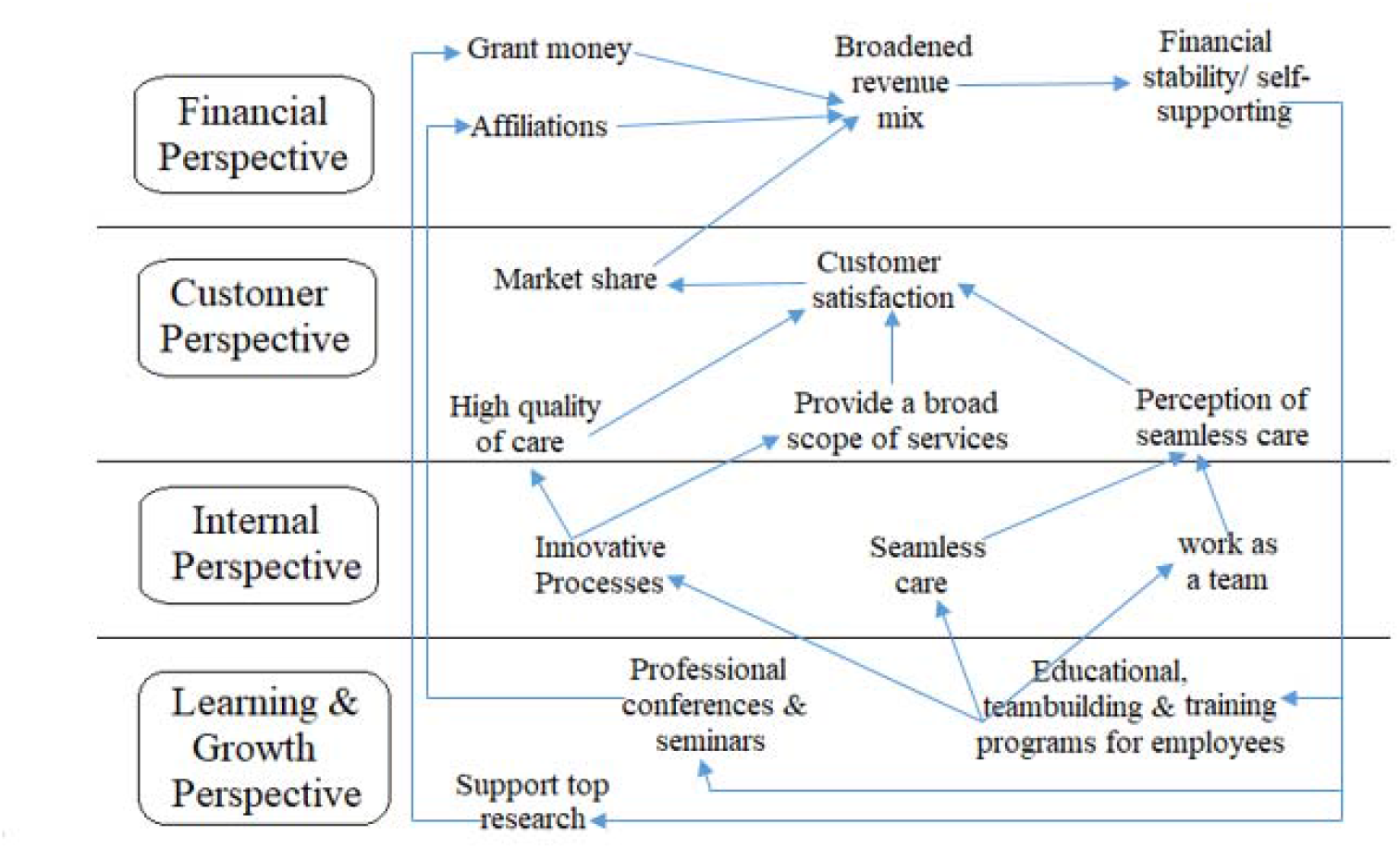
Duke University Health System Strategic Map [26].

Our previous systematic review [27] proves that BSC implementations were effective in improving the financial performance of HCO, elevating patient satisfaction rates, and to a lesser extent improving HCW satisfaction rates. Another review [28] revealed that there had been a lack of engaging stakeholders in BSC implementations, such as engaging patients and HCWs. However, researchers have pointed to the importance of patient and HCW engagement in the process of PE and delivery improvement [29–31]. The rest of the BSC reviews [28, 32–42] focused only on the general narration of the BSC perspectives and subdimensions used. Moreover, none of them summarized the perspectives or dimensions of BSC based on their importance or frequency of use by health care managers. In other words, all the previous systematic reviews lack a systematic methodological categorization of perspectives, dimensions, and KPIs.

In correspondence with this research gap, this review aims at a) finding and recategorizing all the perspectives, dimensions, and KPIs that were employed in BSC implementations for unification purposes, b) ranking dimensions according to their frequency of use by HCO worldwide, and c) ranking dimensions according to their importance from the health care managers perspective.

## Methods

This systematic review is part of broad research. After assessing the impact of the BSC on stakeholder satisfaction [27] and before developing instruments to engage stakeholders in BSC implementations, we sought to accomplish the previously mentioned aims to summarize which dimensions were the most frequently used and essential as per health care managers in implementing the BSC. This review was conducted according to the 27-point checklist of the Preferred Reporting Items for Systematic Reviews and Meta-Analyses (PRISMA) checklist [43], see Appendix (S1).

### 2.1. Eligibility criteria

The inclusion and exclusion criteria were set as shown in Table (1). Table (1) is to be placed here.

**Table 1.** Inclusion/Exclusion Criteria and Search Strategy for PubMed.

| **Table (1)**<br>**Inclusion/Exclusion Criteria and Search Strategy for PubMed** |  |  |  |
| --- | --- | --- | --- |
|  | **Inclusion criteria** | **Exclusion criteria** | **Search Strategy (MeSH terms and keywords) for PubMed** |
| **Population** | Health care organization which offers a primary, secondary, or tertiary health care or medical services such as (clinics, entire hospitals, or hospital's department), without restriction to the ownership or administrative type. | Laboratories, pharmacies, biobanks, radiology departments, hospice homes and medical education centers, unless they were department or unit in the previously included institution types. | hospitals[MeSH Terms]<br>hospital department[MeSH Terms]<br>health[MeSH Terms] |
| **Intervention** | Performance assessment of health care organizations through explicitly implementing BSC, or implicitly assessing the perspectives described in the initial BSC design [6]. | Studies which explicitly used other TQM tools such as the MBNQA, ISO, SQA, six-sigma, etc. | "quality indicators, health care"[MeSH Terms]<br>scorecard*[Text Word]<br>"score card"*[Text Word] |
| **Outcome** | Full reporting of indicators measurements or values. | No reporting or partially reporting of indicators measurements or values. | patient satisfaction[MeSH Terms]<br>cost-benefit analysis[MeSH Terms]<br>health care costs[MeSH Terms]<br>Hospital personnel management[MeSH Terms]<br>staff development[MeSH Terms]<br>knowledge management[MeSH Terms]<br>efficiency, organizational[MeSH Terms] |
| **Study design** | All study designs. | — | No limitation regarding study design, type or time was set in the search strategy. |

Balanced Scorecard, BSC; Total Quality Management, TQM; Malcolm Baldrige National Quality Award, MBNQA; International Organization for Standardization, ISO; Singapore Quality Award, SQA;

### 2.2. Data sources, search strategy, and study selection

First, a search strategy was developed for the PubMed database (see Table 1). Then, the strategy design was adapted for the Embase, Cochrane CENTRAL, and Google Scholar databases. Furthermore, an additional search in the Google engine was performed to find gray literature or unpublished papers, including theses and conference abstracts. Additionally, the reference lists of all the eligible articles were checked by the first author. The databases were searched until October 2020.

The search strategy was developed by the first, second, and fourth authors; the first two were experts in health care management and BSC, while the fourth author was an expert in systematic reviews and meta-analysis. It was initially developed for the PubMed database based on the PICO (Population, Intervention, Comparison, and Outcome) tool [44]. Moreover, the search strategy was developed to include both Medical Subject Headings (MeSH) terms and keywords to widen the search frame and obtain more results. Then, appropriate truncation and relevant indexing terms were used. See Appendix (S2), which shows search strategies in all databases. Afterward, the first author conducted an electronic database search and removed duplicates using the EndNote X9.2 program.

The selection of eligible studies was performed independently by the first and second authors in all steps. Disagreements were resolved by discussion after each step or, if necessary, through arbitration by the fourth author. First, the articles’ titles and abstracts were examined to eliminate irrelevant papers between November 2020 and February 2021. Then, full texts were carefully inspected to decide the final papers’ inclusion list between February and June 2021. If different KPIs were used in more than one implementation in the same study, each was counted as a different implementation. In comparison, implementations using the same KPIs in other locations or times at the same research were considered one implementation. Authors of studies with no available full texts or with partially reported results were contacted for missing data.

### 2.3 Data collection process

Data extraction was performed by the first and second authors independently between June and July 2021 and then compared to discuss differences. The following data were extracted from the eligible studies: 1) author/s, 2) year of publication, 3) country of origin, 4) data collection duration, 5) data collection tool, 6) the number of perspectives, 7) the number of KPIs, 8) availability of weights/importance for perspectives or KPIs, and 9) outcome, which is represented in the KPIs that have been used and their weights/importance. The frequency of each KPI used at each implementation was plotted on Microsoft Excel, and the sum was calculated. In addition, the weight/importance assigned for each KPI at each implementation was reported on a scale of 100%. In the case of studies that did not give weights/importance explicitly, each KPI weight/importance was calculated by dividing one at the number of used KPIs in that study to assign an equal weight/importance for each KPI.

Consequently, the first and second authors independently computed an average of the weights/importance assigned for each KPI. Next, the first author performed regrouping and coding for the KPIs to find the frequency of use and the set weights/importance percentages for each dimension. Then, the resulting major and subdimensions were listed and described between August and September 2021.

The research design of eligible studies was extracted directly from the studies. However, if the research design was not explicitly mentioned, the first and second authors determined it based on the role of the investigator in that study. Specifically, the study was considered observational if the BSC exposures were naturally determined, and the investigator had no part. On the other hand, the study was considered experimental if the investigator actively assigned the BSC intervention.

### 2.4 Quality assessment

The risk of bias (RoB) assessment was performed by the first and second authors independently between August and September 2021 to assess the quality of the included studies. The Risk of Bias in Non-Randomized Intervention Studies (ROBINS-I) tool was used to evaluate the observational and quasi-experimental studies [45]. In comparison, the Cochrane (RoB 2) tool was used for the assessment of randomized controlled trials (RCTs), as per the Cochrane collaboration’s guidelines [46]. The RoB was analyzed at the study level and across studies since authors should avoid summarizing the overall RoB as per the Cochrane Handbook guidelines [47, 48].

In the RoB 2 tool, five types of bias were assessed: bias arising from the randomization processes, bias due to deviations from intended interventions, bias due to missing outcome data, bias in the measurement of outcomes, and bias in the selection of the reported results. On the other hand, in the ROBINS-I tool, seven types of bias were assessed: bias due to confounding, bias in the selection of participants in a study, bias in measurement/classification of interventions/exposures, bias due to deviations from intended interventions/exposures, bias due to missing data, bias in the measurement of the outcomes, and bias in the selection of the reported results.

Using the RoB 2 tool, each type of bias was assessed as low, high, or unclear. While using the ROBINS-I tool, each type of bias was evaluated into five categories: low, moderate, serious, critical, or no information. Afterward, the assessment results of the two reviewers were compared. In case of disagreement, the fifth and sixth authors were consulted. Figures for RoB were prepared using the ROBVIS (Risk Of Bias VISualization) tool [49]. Last, it was recommended not to advocate quality appraisal as a criterion for inclusion in reviews [50]. Therefore, the authors decided to include all studies in this systematic review regardless of their quality assessment.

## 3. Results

### 3.1 Study selection

A total of 4028 studies resulted from running the search strategy in the four databases. In addition, another three studies were identified through a Google search. Therefore, a total of 4031 studies have resulted. Duplicates were removed (n=1046) using the EndNote program, and then the remaining articles were screened based on their titles and abstracts (n=2985). Irrelevant papers were excluded (n=2794).

Consequently, the remaining 191 studies were examined by reading the full texts. Among these papers, 22 papers were written in non-English languages, including Spanish, German, French, Chinese, and Persian. A full-text translation was performed for each study to decide whether to include or exclude any of them. As a result of reading the full texts, 158 studies were excluded, and only 33 were eligible for this review, in which 36 full implementations of different BSC designs were actually applied. Table (2) shows a summary of the 36 implementations. Details of the study selection process are shown in the PRISMA flowchart [43]. See Figure (3) above.

**Figure 3:**
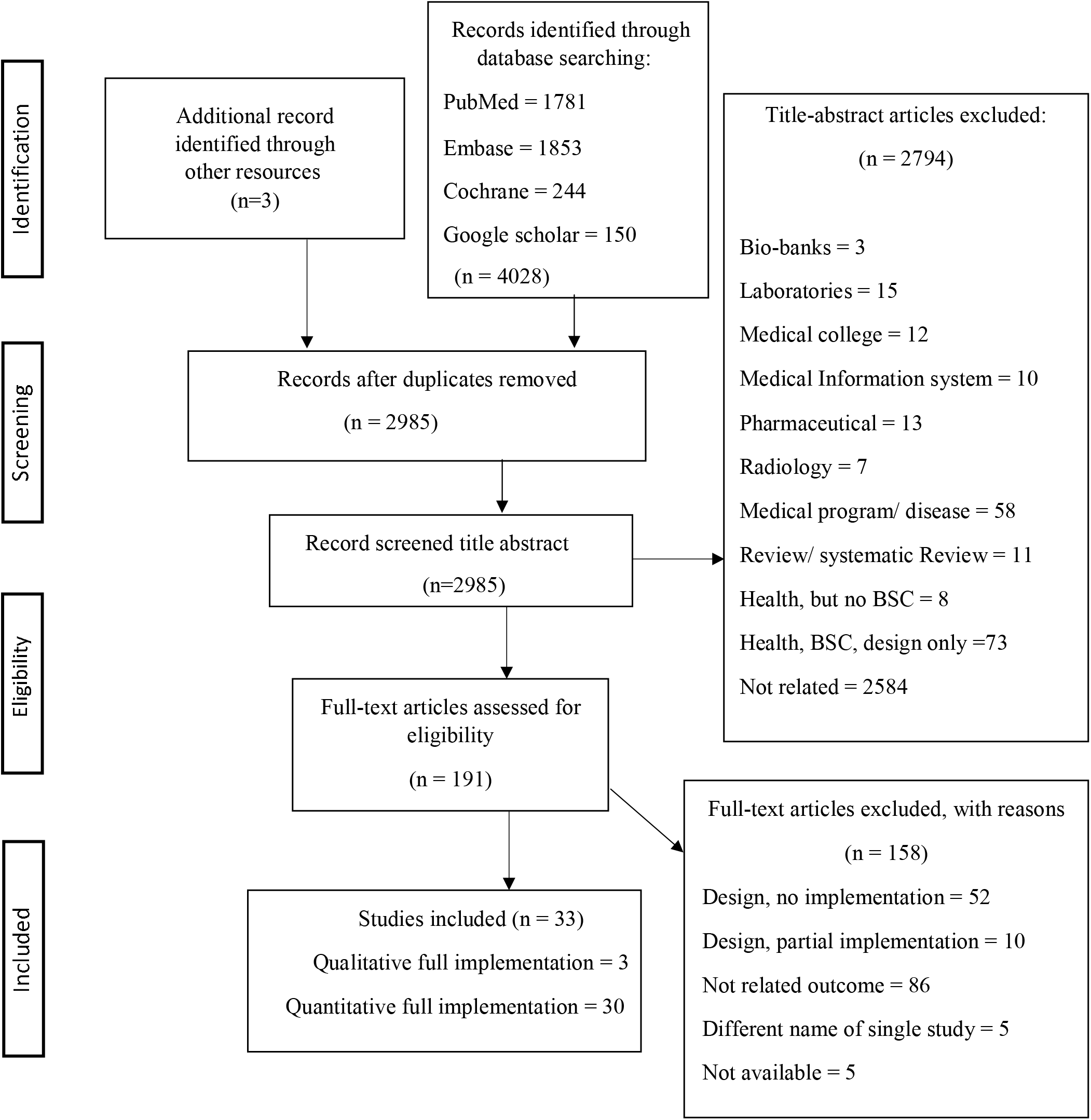
PRISMA Flow Diagram.

**Table 2:** Overview of included studies.

| **Table (2): Overview of included studies** |  |  |  |  |  |  |  |  |
| --- | --- | --- | --- | --- | --- | --- | --- | --- |
| **Author(s)** | **Year of publication** | **Country** | **Duration of data collection** | **Health organization type** | **Data collection tool** | **Number of perspectives** | **Number of indicators** | **Weight /importance (Yes/No)** |
| Pink et al.[69] | 2000 | Canada | 1997-1998 | One hospital | Surveys + hospital reports | 4 | 38 | No |
| Zbinden et al. [76] | 2002 | Switzerland | April- October 2001 | Three departments at a hospital | Personnel statistics and management system + annual reports + questionnaires + accounting system | 4 | 18 | Yes |
| Griffith & Alexander [70] | 2002 | The USA | 1996-1998 | 2300 community hospitals | Medicare database | 4 | 9 | No |
| Biro et al. [71] | 2003 | The USA | 1998-2001 | 63 centers and clinics | Chart audits + surveys + hospital data | 5 | 17 | Yes |
| Smith & Kim [72] | 2005 | The USA | 2001-2004 | Two departments in two hospitals | Survey + audit checklists | 5 | 24 | No |
| Devitt et al. [73] | 2005 | Canada | 2004-2005 | One hospital | Hospital records | 5 | 26 | No |
| Martinez-Pillado et. al. [51] | 2006 | Spain | 2005 | One hospital | Hospital records | 5 | 32 | No |
| Goodspeed [74] | 2006 | The USA | January 2006 | One hospital | NR | 4 | 17 | No |
| Yang & Tung [53] | 2006 | Taiwan | 2000-2002 | 21 hospitals | Secondary data from the department of health + questionnaires | 4 | 16 | No |
| Chen et al. [54] | 2006 | China & Japan | In Japan (April 2003- March 2004). In China (January 2003- December 2003) | Two hospitals | Hospital measurement model | 4 | 19 | No |
| Peters et al. [61] | 2007 | Afghanistan | January- October 2004 | 617 health facility | National Health Services Performance Assessment + patient interviews + HCW & community members | 6 | 29 | No |
| Josey & Kim [75] | 2008 | The USA | December 2006 | One hospital | HCW satisfaction survey + Gallup for patient satisfaction | 5 | 26 | No |
| Chang et al. [86] | 2008 | Taiwan | 2001-2005 | One hospital | NR | 5 | 12 | No |
| Hansen et al. [63] | 2008 | Afghanistan | 2004-2006 | >600 health facility | National Health Services Performance Assessment + patient and HCW interviews | 6 | 29 | No |
| Chu & Wang [64] | 2009 | Taiwan | 2004-2006 | One department at a hospital | Data extraction from hospital financial and performance records + questionnaire to director, assistant directors, head nurses & supervisors | 4 | 11 | Yes |
| Lupi et al. (1) [78] | 2011 | Italy | 2007- 2009 | One hospital unit | Data extraction from hospital records | 4 | 26 | Yes |
| Lupi et al. (2) [78] |  |  | 2008-2009 |  |  | 4 | 34 | Yes |
| Edward et al. [65] | 2011 | Afghanistan | 2004-2008 | 615 health facilities | Performance Assessment + National Health Services patient and HCW interviews | 6 | 29 | No |
| Chen et al.[66] | 2012 | Taiwan | 2004-2010 | 67 departments at a medical center | Secondary data collected by repeated measurements | 4 | 9 | Yes |
| Khan et al. [67] | 2013 | Bangladesh | January-February 2009 | 637 Health facilities | Questionnaire and exit interview questionnaire for clients | 4 | 19 | No |
| Lin et al. [68] | 2013 | China | July 2008-December 2009 | One hospital unit | NR | 4 | 32 | Yes |
| Ajami et al. [55] | 2013 | Iran | NR | One hospital department | Top managers interview questionnaires + staff observations | 4 | 20 | No |
| Mutale et al. [80] | 2014 | Zambia | 2011-2013 | 12 health facilities | HCW & patient Interviews + patient observations + households survey | 7 | 20 | No |
| Rowe et al. [56] | 2014 | Afghanistan | March- August 2010 | 24 health facilities | Patient-provider clinical interactions observations+ Patient follow-up exit interviews + HCW interviews + facility record audits | 5 | 26 | No |
| Edward et al. (1) [57] | 2015 | Afghanistan | 2012 | One health facility | Quantitative and qualitative community survey | 2 | 19 | No |
| Edward et al. (2) [57] |  |  |  |  |  | 2 | 16 | No |
| Edward et al. (3) [57] |  |  |  |  |  | 2 | 17 | No |
| Rabbani et al. [58] | 2015 | Pakistan | 2012 | Six health centers | Survey + services assessment + patient questionnaire exits interviews + HCW questionnaire interview | 5 | 20 | No |
| Teklehaimanot et al. [81] | 2016 | Ethiopia | January – February 2010 | 433 health facilities | Structured & semi-structured internationally accepted questionnaires (health facility audit + HCW interviews, community interviews) | 6 | 32 | No |
| Catuogno et al. [77] | 2017 | Italy | 2007-2008 & 2014 -2015 | One department at a hospital | Stakeholder satisfaction questionnaires + hospital discharge report + charity report + departmental report + hospital discharge database | 4 | 25 | No |
| Gao et al. [59] | 2018 | China | NR | Five hospitals | HCW questionnaires + Patient interview-based questionnaire + TOPSIS method | 4 | 36 | Yes |
| Ebrahimpour et. Al [52] | 2019 | Iran | 2010-2017 | One hospital | Hospital records | 4 | 23 | No |
| Widyasari & Adi [60] | 2019 | Indonesia | During 2018 | One hospital | structured interviews + semi-structured interviews + documentation + Observation | 4 | 11 | Yes |
| Mabuchi et al. [82] | 2020 | Nigeria | April-May, 2016 | 111 primary health facilities | Survey + interview questionnaire | 6 | 32 | No |
| Manolitzas et. al. [79] | 2020 | Greece | NR | One hospital department | Interviews + hospital records + observation | 4 | 11 | Yes |
| Gonzales et. al. [83] | 2020 | NR | NR (but data extracted 2018-2019) | One medical center | Hospital records | 8 | 13 | No |
|  |  |  |  |  |  | <u>Average</u><br>4.5 | <u>Average</u><br>22 | <u>Total</u><br>Yes: 10<br>No: 26 |

United States of America, USA; Health Care Workers, HCW; Technique for Order of Preference by Similarity to Ideal Solution, TOPSIS; Not Reported, NR;

### 3.2 Study characteristics

#### 3.2.1 Language and location

From the resulting 36 implementations, one was in Spanish [51], one was in Persian [52], and the rest were in English. The 36 implementations were performed in various countries: 19 in Asia [52–68], seven in North America [69–75], six in Europe [51, 76–79], three in Africa [80–82], and one without location information [83].

#### 3.2.2 Settings

Twenty-one implementations were performed in hospitals (secondary and tertiary HCO) [41, 51–55, 60, 61, 63, 65, 69, 70, 72–79] and 15 in medical centers or health facilities (primary HCO) [56–58, 62, 64, 66–68, 71, 80–83].

#### 3.2.3 Implementations

Few studies had more than one implementation; [57, 78] included three and two implementations, respectively, with different KPIs per implementation. Thus, the 33 resulting studies contained 36 unique implementations. No BSC implementation in the COVID-19 era was found.

#### 3.2.4 Study design

The 36 BSC implementations varied in their designs. However, most studies did not explicitly report their study design. We categorized the 36 implementations based on the active role of the investigator in BSC implementation and the time of data collection. Consequently, one sole study design was RCT [80]. Moreover, 14 implementation designs were uncontrolled quasi-experiments. Specifically, six implementations had a posttest-only design [54, 55, 59, 76, 79, 82]. Five implementations in four studies had pretest-posttest designs [52, 61, 77, 78]. Finally, three implementations interrupted the time series design [65, 71, 84]. On the other hand, 20 implementations were observational; six implementations in four studies were cross-sectional [56–58, 70], one implementation was prospective [85], ten implementations were retrospective [53, 62–64, 66–68, 74, 75, 81], and two implementations were prospective and retrospective [72, 73]. Finally, one implementation did not have sufficient information or reported the study design [83].

### 3.3 Decision model

Some of the resulting studies integrated multiple-criteria decision analysis (MCDA) with BSC. One study [79] combined BSC with simulation and MCDA techniques with what was referred to as S-MEDUTA. Another study [54] integrated the BSC with fuzzy analysis. Two studies [59, 65] combined BSC with AHP, and one [59] used the TOPSIS technique. Studies explained that using these methodologies with the BSC would help them arrive at more informed and better decisions.

### 3.4 Perspectives frequency of use and importance

A total of 797 KPIs were extracted from the resulting implementations. These KPIs were categorized in the studies under 15 perspectives. The average number of perspectives used per study was 4.5, and for the KPIs, it was 22. The most frequently used perspectives were the internal, financial, patient, learning and growth, HCW, managerial, community, and stakeholder perspectives. The total use frequencies of these perspectives at the implementations were 29.6%, 17%, 12.6%, 12.6%, 9.4%, 6.3%, 5%, and 3.1%, respectively. On the other hand, the topmost important perspectives from the health managers’ viewpoint were the internal, financial, learning and growth, patient, HCW, community, managerial, and stakeholder perspectives with a total weight/importance of 37.9%, 15.4%, 12%, 11.3%, 7.8%, 7.7%, 3.6%, and 2.8%, respectively.

### 3.5 Categorization and regrouping of KPIs

The 797 extracted KPIs were plotted according to their frequencies and weights/importance in the categorization process. After regrouping these KPIs into homogenous major dimensions and subdimensions, 13 major dimensions resulted, with 45 subdimensions. The resulting major and subdimensions are listed and described in Table (3).

**Table 3:** Description of the BSC Major- and Sub-dimensions.

| Table (3): Description of the BSC Major- and Sub-dimensions |  |  |  |
| --- | --- | --- | --- |
|  | The major-dimensions | Description | Description of sub-dimensions and their KPIs: |
| 1 | **Financial dimension** | Represented as the financial perspective in BSC.<br>It consisted mainly of four sub-dimensions. | **The first sub-dimension:**<br>Margins such as the cash flow margin and the operating profit margin.<br>**The second sub-dimension:**<br>Expenditures and costs such as personnel costs, controllable costs, and the cost per case or admission.<br>**The third sub-dimension:**<br>Revenues as the revenue per admission and the return per employee.<br>**The fourth sub-dimension:**<br>Revenues versus expenditures ratios, such as, ROI, ROA, and capital turnover. |
| 2 | **Error-free and safety dimension** | Part of the internal perspective of BSC. It constituted of five sub-dimensions | **The first sub-dimension:**<br>Mortality, such as net death rate per 1000 patients or gross mortality.<br>**The second sub-dimension:**<br>Errors, accidents and complications sub-dimension, which contained KPIs such as complications index, hospitalized accident rate, medication error rate, blood preparation error, pneumonia complications.<br>**The third sub-dimension:**<br>IC index such as postoperative infection rate and infection prevention. |
|  |  |  | **The fourth sub-dimension:**<br>HW management includes segregating waste into proper sharps, infectious, pathological, pharmaceutical, radioactive and non-hazardous waste disposals. As well as waste minimization, color-coding, labelling, handling, transports, storage, treatment and disposal.<br>**The fifth sub-dimension:**<br>Safety standards focused on patient safety through the appropriate efforts to avoid adverse events related to errors in diagnosis, medication, or treatment, such as the full implementation of the 27 Safe Practices for Better Health care standards and percentage implementation of the National Patient Safety Goals. |
| 3 | **Efficiency and effectiveness dimension** | Part of the internal perspective. Through this dimension, four sub-dimensions are mainly focused on. | **The first sub-dimension:**<br>Number of admissions, visits and diseases; which evaluated the number of patients, surgeries, admissions, re-admissions, cross-appointments, and disease scores.<br>**The second sub-dimension:**<br>Efficiency, utilization, and productivity included KPIs such as the case mix index, productivity percent, service utilization, ER patients per year per doctor, admitted inpatients per year and doctor, bed turnover rate, and nurses' workload.<br>**The third sub-dimension:**<br>The improvement sub-dimension, such as the cure, recovery, and improvement rates.<br>**The fourth sub-dimension:**<br>The occupancy, which mainly focused on bed occupancy rate, indicates the percentage of beds occupied by patients in a given period |
| 4 | **Availability and quality of supplies and services dimension** | Part of the BSC internal perspective. Under this dimension, three sub-dimensions are mainly focused on. | **The first sub-dimension:**<br>Medications, which included drug management, drug availability index, and tracer drug index.<br>**The second sub-dimension:**<br>Supplies and equipment, such as supply distribution system and equipment functionality index.<br>**The third sub-dimension:**<br>Products and services, this sub-dimension included KPIs, which evaluate either the variety of medical services or products offered by the HCO or their quality. |
| 5 | **Time dimension** | Part of the BSC internal perspective. It constituted of three sub-dimensions. | **The first sub-dimension:**<br>The operation processing time is the time needed from the initiation of service until completion, for example, billing time, treatment time, correcting mistakes time, retrieving an archived file time, etc.<br>**The second sub-dimension:**<br>WT or the delay time until providing services is initiated.<br>**The third sub-dimension:**<br>LOS of an admitted patient till discharge. |
| 6 | **HCO building dimension** | It is usually part of the internal or the customer perspectives.<br>It included one sub-dimensions | Constituted of one sub-dimension, it included KPIs which are related to HCO's building. For example, HCO's capacity, ER volume, waiting area, bathroom, cleanliness, water, electricity, appointments, ease of access, and ambulance availability. |
| 7 | **The responsiveness and communication dimension** | It is usually part of the customer perspective in BSC.<br>It constituted of three sub-dimensions. | **The first sub-dimension:**<br>Response to patients' needs, including response to their inquiries and feedback. This was performed either after patient arrival, during the admission and the treatment process, or before the discharge.<br>**The second sub-dimension:**<br>Patient information including patient information, education, guidelines, counselling and consultation services.<br>**The third sub-dimension:**<br>Communication which included evaluation for the nature of both internal and external communications, for example, the ability of coordination and teamwork among HCWs, and the relationships between HCWs and patients. |
| 8 | **Patient-centeredness dimension** | It is usually evaluated as a part of BSC customer or the stakeholder perspectives.<br>It included focusing on three sub-dimensions. | **The first sub-dimension:**<br>Complaints which mainly concerned with measuring the patients' complaint rate.<br>**The second sub-dimension:**<br>Patient satisfaction focused on measuring patient satisfaction rate in general or the satisfaction rate in specific toward a medical service, a specific HCWs dimension, or in a particular HCO department.<br>**The third sub-dimension:**<br>Patient loyalty is usually measured by patient retention and recommendations for that HCO. |
| 9 | **The HCW-centeredness dimension** | Either a part of the customer or the internal perspectives at BSC, while some studies added it under HCWs management perspective [30,48,55,62,67,72]. It consisted of six sub-dimensions. | **The first sub-dimension:**<br>The staffing process starts from the employment to the introduction process of the new employees.<br>**The second sub-dimension:**<br>HCWs' engagement and motivation include engaging doctors and nurses in the HCO managerial decisions and plans, high-performing HCWs rewarding, upgrade rate on the career ladder, involvement in bonus decision, and evaluation of HCWs motivation and burnout rates.<br>**The third sub-dimension:**<br>HCWs feedback, for example, HCWs perception index surveys.<br>**The fourth sub-dimension:**<br>HCWs' satisfaction by doctors', nurses', other HCWs' satisfaction rates evaluation.<br>**The fifth sub-dimension:**<br>HCWS' loyalty index includes the doctors' and nurses' willingness to stay at the same HCO for another five years and recommend their colleagues to work at their HCO.<br>**The sixth sub-dimension:**<br>HCWs' turnover which assessed the number of doctors, nurses, etc. who left their jobs at that HCO in a specific period. |
| 10 | **The HCWs' scientific development dimension** | Part of the innovation and knowledge, or the customer perspectives.<br>It constituted of three sub-dimensions. | **The first sub-dimension:**<br>HCWs' KAP, which concentrated on the HCWs' current competencies and knowledge in different medical and health related fields, the percentage of skillful employees, and their attitudes, behavior, and punctuality.<br>**The second sub-dimension:** |
|  |  |  | HCWs' training, through which the number of seminars, courses performed to improve the HCWs' KAP and the budget specified for this purpose were evaluated.<br>**The third sub-dimension:**<br>Research and scientific productivity, which can be a result when the previous two sub-dimensions are improved. For example, impact factor per HCW, total research impact of the HCO, number of participations in conferences and research programs per year, and the expenditure on the medical research. |
| 11 | **The technology and information system dimension** | Part of the innovation and knowledge perspective.<br>It consists of three sub-dimensions. | **The first sub-dimension:**<br>Records, such as: patient record index, HMIS records, hospital laboratory registrations, and medical records completion rate.<br>**The second sub-dimension:**<br>Reports, which included the ability to produce reports for different purposes in different HCO departments.<br>**The third sub-dimension:**<br>TI system, which was reflected by assessing the HMIS effectiveness, and the intensity of information use. |
| 12 | **The community and reputation (external) dimension** | Part of the customer perspective, while some studies [48,49,69,72,78] added it under the community and social perspective.<br>It consists of three sub-dimensions. | **The first sub-dimension:**<br>Market share evaluation for the HCO in general or for a specific department at that HCO.<br>**The second sub-dimension:**<br>The CSR focused on the exemptions offered by the HCO for poor patients, benefits provided to the community, teaching and training programs offered for medical students, and the community satisfaction rate.<br>**The third sub-dimension:**<br>Privacy and female considerations evaluated the percentage of female patients, the availability of female doctors and nurses in the HCO, and the patients' privacy adherence. Although the female consideration in HCOs may not have significant importance in all cultures. However, it was vitally considered in some implementations [30,31,48,68,75]. |
| 13 | **The managerial tasks and PE dimension** | Part of the internal perspective. However, some studies added it under the managerial perspective [72,73].<br>It constitutes four sub-dimensions. | **The first sub-dimension:**<br>Standards and regulations, which can be reflected by the standardization capability for different HCO working processes. Moreover, the HCWs' awareness of these standards and regulations highlights the importance of the rules and standards to be clear, understandable, and specific to them.<br>**The second sub-dimension:**<br>Planning and targets sub-dimension incorporated business plan and target setting, updating, and HCWs' awareness and attention to them.<br>**The third sub-dimension:**<br>Internal assessment sub-dimension using managerial quality tools for quality enhancement, managing objectives, or the PE. For example, the included studies assessed the implementation of a continuous<br>quality improvement system, MBO, TQIP, QOPI, and BSC, or the scores resulted from using different instruments and scales such as Press Ganey. This dimension also includes the internal PE process, such as regular performance review meetings, the visualization of performance data, and the HCWs' attention to performance.<br>**The fourth sub-dimension:**<br>The accreditation, peer-reviews represent external assessments and certificates the HCO receives from external sources. It includes evaluating the accreditation status of the HCO, such as JCI or AACI, plans to maintain it, and the performed periodic revising for the accreditation manuals. It also includes the certificates the HCO received, such as the ISO certificate and the peer reviews. |

BSC, Balanced Scorecard; KPIs, Key Performance Indicators; ROI, Return On Investment; ROA, Return on Assets; IC: Infection Control; HW, Health Waste; ER, Emergency Room; WT, Waiting Time; LOS, Length of Stay; HCO, Health Care Organization; HCWs, Health Care Workers; KAP, Knowledge, Attitude, and Practices; HMIS, Health Management Information System; TI, Technology and information; CSR, Community Social Responsibility; PE, Performance Evaluation, MBO, Management by Objectives; TQIP, Trauma Quality Improvement Program; QOPI, Quality Oncology Practice Initiative; JCI, Joint Commission International; AACI, American Accreditation Commission International; ISO, International Organization for Standardization;

### 3.6 Dimensions/Subdimensions frequency of use and importance

The KPIs’ use frequencies and assigned weights/importance at the resulting implementations were plotted separately. Grouping and recategorizing KPIs resulted in Figures (4 & 5) above, showing 13 major dimensions and 45 subdimensions based on their frequency of use and importance, respectively.

**Fig. 4:**
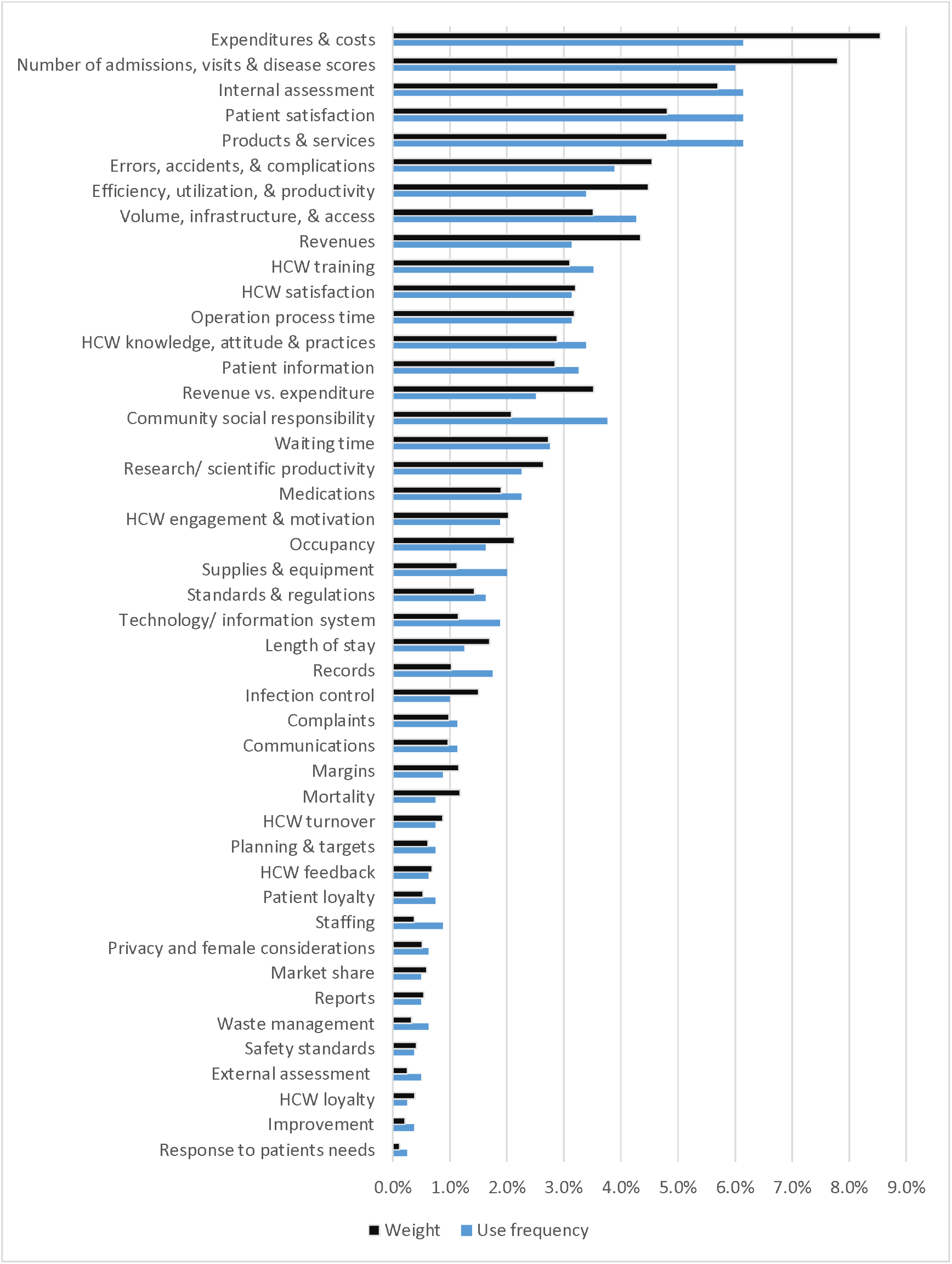
The BSC 45 subdimensions. Figure legend: After re-grouping the 797 indicators, 45 subdimensions resulted. This figure shows the frequency and the weight/ importance for each subdimension independently.

**Figure 5:**
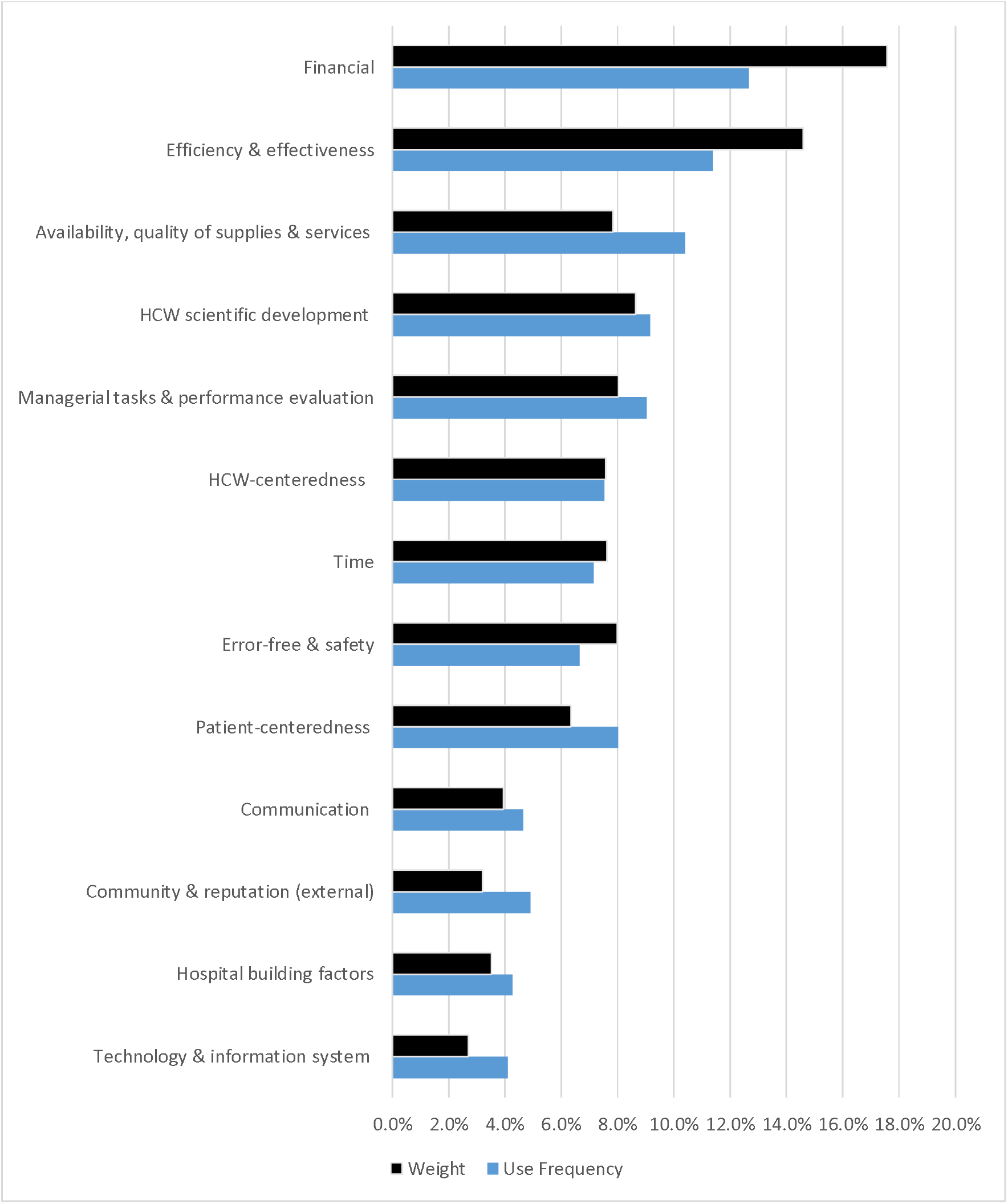
The BSC 13 major dimensions. Figure legend: Reassembling the 45 subdimensions resulted in 13 major dimensions. This figure shows the frequency and the weight/ importance for each major dimension independently.

### 3.7 The quality assessment

Each study was evaluated in terms of RoB, as illustrated in Appendix S3. The RoB 2 tool was utilized to assess the ROB in the sole RCT study [80], for which the assessment showed fair evaluation, except for performance bias. On the other hand, utilizing the ROBINS-I tool for assessing the RoB in observational and quasi-experimental studies revealed no information about confounders’ adjustment methods except in three studies [53, 65, 69]. The confounding agents were apparent in the three studies; one study [69] performed confounders adjustments. On the other hand, another [53] adjusted for patient severity but not for the LOS and mortality rate. Lastly, one study [65] did not perform adjustment at all, which may have affected measurement precision.

The selection bias across studies reflected a serious RoB in five studies [53, 56, 60, 65, 86]. Therefore, the intervention and the follow-up did not coincide together, and a potentially substantial amount of follow-up was missing in their analysis. Studies with a moderate risk of intervention/exposure measurement bias reflected a well-defined intervention status, but some aspects of the assignments of intervention status were determined retrospectively. Furthermore, bias in selecting the reported results was serious in one study that partially reported the results [60]. Studies that reported all results but did not have a preregistered protocol or their outcome measurements were not defined in an initial plan were given a moderate risk. See (S3 Appendix).

## 4. Discussion

### 4.1 Discussion of the main results

All the perspectives, dimensions, and KPIs employed in BSC implementations were collected to fulfill the research aims. Categorization and regrouping of the KPIs into major and subdimensions were performed. Then, the dimensions were ranked according to their frequency of use, as well as their importance. The BSC tool can offer comprehensive planning, monitoring, evaluation, and improvement of HCO KPIs. Hence, their performance should be improved in the short and long term.

In general, studies had either no information or low or moderate ROB. At the same time, only a few of them had serious or critical ROB. However, studies that had only fully reported BSC indicator measurements were included. Many of them did not have a preregistered protocol or predefined measures at their plan. No information was found regarding confounders or deviation from unintended interventions.

### 4.2 Overall completeness and applicability of evidence

Analyzing the results shows that BSC implementations typically utilized four fundamental perspectives: financial, customer, internal, and innovation and knowledge. However, the frequent employment of other BSC perspectives shows the need for slight modifications of BSC design. For example, adding the sustainability, external, environmental, or community perspective focused on the community’s needs, perceptions, and reputation of the HCO. This finding corresponds with a study [141] that referred to the sustainability perspective of the BSC as the fifth pillar. Additionally, there was a need to add the managerial perspective, which included merging the strategy with other BSC perspectives and evaluating administrative tasks and tool utilization at HCO.

The variation among BSC implementations in the categorization of the same KPIs reflects the need for data standardization. HCW training-related KPIs, for example, were categorized under the learning and growth perspective in almost half of the resulting studies [53, 54, 65– 68, 74, 75, 77, 78, 80, 81, 55–59, 62–64]. Meanwhile, the rest of the studies categorized them under the perspectives of HCW [80, 81], quality [71], service capacity, provision/service capacity [56, 62, 63, 66, 68, 81], and healthcare facility functionality [58]. These results are consistent with a study [5] that referred to the lack of defining measures and the lack of data standardization. The differences in categorization prove our assumptions in the calculation imprecision in the previous reviews. Specifically, in the use frequency or the importance of the perspectives and KPIs. Our systematic review solved this calculation bias by uniformly forming the 797 KPI categorizations. Regrouping similar or semisimilar KPIs under the same category resulted in more precise results. Unification of dimensions can guide uniform future implementations of PE or BSC at HCO, allowing data sharing and comparability. Dimension unification can be why our findings are different from another systematic review [142] that did not consider unifying the classification of KPIs. According to HCO management, the average LOS, HAIs, patient satisfaction, bed occupancy, and bed turnover rate were the most useful KPIs.

Analyzing the results also shows a lack of BSC utilization in HCO during the pandemic. Additionally, there has been a lack of studies comprehensively examining the impact of COVID-19 on KPIs. Our analysis reflects that most KPIs were negatively affected during the pandemic, except the IC and safety measures, which improved in some cases. A comprehensive PE of HCO during the COVID-19 pandemic worldwide is required. Some dimensions that are essential for PE are still poorly investigated. Therefore, we recommend that future researchers perform a comprehensive PE for HCO during COVID-19 using the measurements of the resulting dimensions. This analysis will provide a better understanding of the dimensions of the causal relationships between them. It will also allow comparability of the interventions’ outcomes, which will boost the performance and mitigate the consequences of the pandemic on HCO. Moreover, researchers are encouraged to perform systematic reviews for each dimension, especially those that are already well investigated and the investigation of dimensions that are still poorly investigated but essential for PE.

### 4.3 Practical assessment implications of the resulted dimensions at the COVID-19 era

This review can guide healthcare managers and researchers since the resulting dimensions can be utilized to synthesize future BSC measurements. Specifically, the dimensions can direct the creation of new instruments to engage stakeholders in future BSC implementations. Moreover, this review can provide a road map for healthcare managers to perform a comprehensive PE of HCO during the COVID-19 pandemic.

Although this systematic review included ten months after the initiation of the COVID-19 pandemic, no research on BSC utilization in COVID-19 was found. Moreover, health policy experts stated that insufficient standardization of quality measurement approaches in the COVID-19 era challenged the sharing purposes. As a result, the comparison between the performance of healthcare systems is disrupted [5]. Comparison is critical in cases where the optimal performance is not fully understood as in pandemics, and a comparison with other health systems would be informative and necessary [5].

Therefore, addressing the lack of data standardization was suggested to be established by quickly defining measures, which could allow health systems, at least in the short term, to use standardized methods to better understand their performance [5]. The authors pursued further analysis in this paper based on independent studies per resulting dimension during the COVID-19 era to highlight how these dimensions can be utilized to monitor and improve HCO’s performance during the COVID-19 pandemic.

#### 4.3.1 The Financial major dimension

Due to COVID-19 hospitalizations at the beginning of the pandemic, health policy experts suggested that HCO in some regions will have more significant revenue and greater costs related to additional HCW and resources. In contrast, other hospitals will experience mostly sharp reductions in elective and outpatient payments, which will create unprecedented financial challenges for HCO [87]. However, in addition to the higher costs of HCWs and resources, researchers found higher costs of treatment due to extra diagnostic tests and isolation costs [88].

In the United Kingdom (UK), the total expenditure on the National Health System (NHS) has increased significantly during the pandemic [89]. The NHS made funding upgrades to expand waiting areas and treatment cubicles [90]. Some studies have focused on cost-effectiveness calculations. A study in South Africa indicated that purchasing intensive care unit (ICU) capacity from the private sector during COVID-19 surges may not be a cost-effective investment [91]. To date, there is still a lack of studies that handle the financial dimension or developed cost-saving strategies at the health organization level in COVID-19.

#### 4.3.2 The error-free and safety major dimension

This dimension includes monitoring, analyzing, and comparing mortality rates and investigating its determinants in HCO. Although mortality may not be directly related to errors, mortality rates higher than the average can reveal an underlying mistake. A cohort study in Mexico City [84] found that the mortality rates at the hospital’s ICU and non-ICU departments were similar. The reason behind this finding was the ICU bed’s unavailability. Approximately 45% of the patients who did not survive did not receive an ICU bed, which raised the mortality rate in the non-ICU admitted patients. However, this study revealed that the leading cause of non-ICU-admitted patients was acute respiratory distress syndrome (ARDS). The leading cause of mortality for admitted patients was septic shock, followed by ARDS and multiorgan failure.

The WHO has provided clear guidelines for infection control (IC) during healthcare when COVID-19 is suspected or confirmed [92]. Patient safety was investigated in a systematic review of Indian-related studies [93]. Patient safety was negatively impacted during the COVID-19 pandemic due to inadequate preparation of the healthcare system, such as infrastructure and human and material resources. Additionally, researchers categorized diagnostic errors that could occur in the COVID-19 pandemic into eight types and suggested how to reduce them [94].

However, many studies have shown improvements in this dimension during the pandemic. A study in the UK [95] found a significant increase in the Safety Attitude Questionnaire (SAQ) scores of doctors and other clinical HCWs and no change in the nursing group. It also showed a significant decrease in error reporting after the onset of the COVID-19 pandemic. Another study in Iran [96] found that health-associated infections (HAIs) during the pandemic were reduced, which could be referred to as the proper implementation of IC protocols. This finding is supported by a study in Ghana [97], which found that HCW compliance with IC measures was high during the pandemic.

The health waste (HW) management subdimension was intensively investigated due to the tremendous increase in HW volume during the pandemic [98]. A study in Iran [99] indicated that infectious waste increased by 121% compared with before the pandemic. Direct exposure of HCW to virus-contaminated waste with inadequate safety measures and mismanagement of HW may lead to their infection and facilitate the transmission of COVID-19 [98, 100]. The WHO has provided clear guidelines for managing healthcare waste during the pandemic [101]. Despite that, many studies worldwide [100, 102, 103] illustrated the existence of gaps and a flawed system for handling HW during the pandemic.

A mini-review [98] of HW during the pandemic showed that disinfecting waste, followed by proper segregation and on-site treatment, can also provide better and healthier HW management. It also revealed that surplus HW accommodation, mobile treatment, and temporary storage strategies might aid the sustainable management of healthcare waste without further spreading the virus. Another study in Brazil [102] proposed a model for the proper management of HW. It focused not only on the operational management KPIs of the HW but also on environmental management, such as sustainable practices. Moreover, it highlighted the importance of employee training on HW guidelines since HW management was not considered an essential competence or a priority for every HCO.

#### 4.3.3 The efficiency and effectiveness major dimension

Analyzing the number of patient visits and admissions in the USA [104] revealed a decrease in ER visits, increasing hospital admissions. However, another study in Alberta [105] perceived decreased admissions and ER visits to the hospital, despite the low volume of COVID-19 hospital admissions.

Many studies have been performed to analyze the efficiency, utilization, and productivity of HCO during the pandemic. A study [106] indicated that efficient hospitals under normal conditions lost their efficiency during COVID-19 and had to adapt to the new criteria. A systematic review [107] showed that healthcare utilization decreased by approximately one-third during the pandemic, with more significant reductions among people with less severe illnesses.

A study at an isolation hospital in Egypt [108] utilized the DEA tool to improve efficiency. This confirmed that the number of nurses and the number of beds impacted the operational efficiency of COVID-19, while the number of physicians had no significant effect on the efficiency. These results are compatible with a study in Mauritius [19] that found that nurses and beds are the most critical factors in hospital production; that is, a 1% increase in the number of beds and nurses resulted in an increase in hospital outputs by 0.73 and 0.51%, respectively.

#### 4.3.4 The availability and quality of the major dimensions of supplies and services

The supply and logistics management dimension was considered an important KPI in tackling COVID-19 [6]. This dimension includes evaluating the availability and quality of COVID-related medications, masks, personal protective equipment (PPE), detergents, medical services, supportive services, etc. Additionally, researchers viewed the availability of both clinical and supportive services at hospitals as essential in responding to the COVID-19 pandemic and the flow of COVID-positive patients [109]. The spectrum of supportive services to a hospital encompasses linen and laundry, diet, central sterile supply department (CSSD), transport, consumables in large quantities at hospital stores, mortuary, and engineering services [109]. Some of the essential items were filtering face-piece respirators or N95 respirators and the availability of PPE kits [109]. The global challenge during this pandemic in terms of inadequate availability of PPE in HCO highlighted the vital role of CSSD. Centers for Disease Control and Prevention (CDC) suggested a method of decontamination, and reuse of filtering face respirators to overcome the shortage of these respirators is their extended use or reuse [109].

However, researchers referred to the lack of studies on the quality of supplies and services at HCO in COVID-19 [5]. Lack of studies can be referred to as data lag in pandemics: the time between care provision and quality measurement reporting [5]. Policymakers suggested that measures should be less reliant on claims data, which by nature have a time lag, and focus on actions that can be generated from the electronic health record (EHR) [5].

#### 4.3.5 The time major dimension

An “extra layer of processes” was added due to the donning and doffing protocols and cleaning requirements, which slowed all the operational processes down and increased the time required to accomplish serving the medical care to patients [89]. The patient WT was also influenced. In the UK, WT reached high levels in studies with a notable impact on elective surgery. The number of patients who waited for more than a year to receive NHS treatment in July 2020 was 81-fold greater than the previous year’s number [90].

Moreover, patient length of stage (LOS) also increased for another 2–3 days. A reason for that was the delays in COVID-19 testing results [110]. LOS in the USA was two days more than that in Italy and five days less than that in Germany [111]. A systematic review for patient LOS in COVID-19 [112] concluded that LOS in China was longer than that in any other country, referring to differences in criteria for admission and discharge and different timings within the pandemic. Another study [105] found a negative association between the LOS and the case fatality rate. Therefore, LOS estimation can be introduced as a KPI to scale the success of the countries fighting the ongoing pandemic.

Moreover, LOS provides insights into when hospitals will reach capacity and predicts associated HCW or equipment requirements [112]. Discharge status should be considered when analyzing LOS since the patients who were discharged alive have longer LOS than those who died during their admission [112]. Hospitals reported that health insurance plans resisted paying for additional patient days in the hospital while awaiting COVID-19 test results [110].

However, complying with the CDC guidance on testing and disposition of patients was suggested to reduce the patient LOS, freeing up hospital beds for incoming COVID-19 patients [110]. Another study in the UK [110] indicated that due to the complexity and partiality of different data sources and the rapidly evolving nature of the COVID-19 pandemic, it is most recommended to use multiple LOS analysis method approaches on various datasets.

A combination of an accelerated failure time (AFT) survival model and a truncation corrected (TC) method with the multistate (MS) survival model was found to be helpful in epidemic planning and management. Finally, the findings of a cohort study [113] concluded that a multimechanism approach (MMA) effectively decreased the average LOS in the ICU by 5.4 days and up to nine days in older patients. This finding suggests that implementing this treatment protocol could allow a healthcare system to manage 60% more COVID-19 patients with the same number of ICU beds.

#### 4.3.6 The HCO building major dimension

Design and infrastructure preparation were considered essential dimensions in some HCO during the pandemic [6]. Healthcare systems made adaptations in HCO buildings after the COVID-19 pandemic. Examples include expanding waiting areas, increasing ICU capacity, establishing isolation areas, and building new hospitals [90]. In the UK, the NHS temporarily used private hospitals to provide public care, increasing the number of beds, ventilators, and all HCW categories. Moreover, nonhospital sites were temporarily turned into hospitals [114]. However, researchers did not sufficiently investigate the ease of access to HCO during the pandemic.

#### 4.3.7 The responsiveness and communication major dimension

The main goal of HCO was considered to provide high-quality care to patients and meet their needs and expectations during an outbreak such as COVID 19 [106]. Moreover, dialog and listening to health demands in COVID-19-suspected patients was highlighted as the foremost step in the flows of care and guidance [115].

Communication among HCW was also highlighted. A study [4] considered HCW reception of family support, colleagues, support, clear communication, and COVID-19 information as the most valuable resources in the pandemic. Lower HCW psychological distress symptoms, burnout, and intentions to quit were perceived when these communication resources were more available. Another study [116] indicated that gratitude in communication could reduce depression in HCWs by promoting social support and hope.

In addition, communication between HCWs and patients was also investigated during the pandemic. A study in Jordan [117] found that physician–patient communication (PPC) positively affected the patient’s psychological status in COVID-19. It recommended avoiding communication errors using jargon, not being available to patients, and not showing empathy in communication. Additionally, it emphasized the benefit of physicians as excellent listeners to patients. However, HCW-patient communication faced few obstacles during the pandemic. The protective equipment used by HCWs in the pandemic could have imposed a barrier to effective communication or eye contact with them [118]. Some pediatricians reported difficulty communicating with families and following up with patients, especially newly discharged neonates and infants, using the telephone [119].

However, more research is still needed to improve and evaluate patient education programs, patient guidelines, counseling and consultation services, and communication skills between HCWs and patients during the pandemic.

#### 4.3.8 The patient-centeredness major dimension

Many studies have been conducted to evaluate patient satisfaction. A study [120] indicated no difference in patient satisfaction of the period spent in the emergency room before and during the pandemic. Another study [121] showed positive patient experience and satisfaction rates in Saudi Arabia’s largest institutions during the pandemic. Moreover, many studies have focused on the psychological assessment of the impact of COVID-19 on the population in general. However, few studies have focused on specifically assessing the psychological effects on patients. For example, a study [122] found that COVID-19 patients with low education levels and females who have undergone divorce or bereavement tended to have a high prevalence of adverse psychological events. Another study [123] found that the psychological consequences of the pandemic were better handled by cancer patients 65 years of age or older, while younger cancer patients were more psychologically affected. Early psychological status identification and intervention should be conducted to avoid extreme events such as self-mutilating or suicidal impulsivity for patients [122]. The patient complaints and loyalty assessment during the pandemic and the psychological impact of COVID-19 on non-COVID-19 patients still need more investigation.

#### 4.3.9 The HCW-centeredness major dimension

Physicians referred to the importance of reliable acknowledgment and motivation both emotionally and financially, considering the sacrifices they provide every day [119]. Parallel to this, staffing and recruitment of an adequate number of medical and nonmedical HCWs were considered important KPIs for the PE of HCO at COVID-19 [6]. In the UK, the NHS employed strategies to facilitate the staffing process due to the shortage of HCWs. First, newly qualified/final year medicine and nursing students were deployed. Second, the return of the former HCW was made [114].

The HCW satisfaction rate and burnout have been evaluated in many studies during the pandemic. A study [119] showed that the physicians’ burnout prevalence was found to be 57.7% in the pandemic, which is considered so high. HCWs who lack PPE reported lower occupational satisfaction than those who did not [119, 124]. HCW accomplishments during the pandemic were positively associated with higher occupational satisfaction rates [124]. Therefore, emphasizing HCW accomplishments leads to increased satisfaction rates.

Moreover, as mentioned earlier, the better the performance of the communication dimension, including psychological support, will raise HCW job satisfaction and lower the rates of burnout and stress [116, 124]. Some HCWs felt anxiety and fear mainly due to the possibility of transmitting the virus to their family members and the elderly living in their house [119]. A study in Canada [4] showed that HCW training and counseling services were perceived as helpful in reducing HCW stress. Despite that, they were underutilized in HCO.

On the other hand, although most nurses had to increase their workload due to staff shortages, a study [124] found that the elevation of the workload was not associated with lower occupational satisfaction. Additionally, another study in Singapore [125] found that HCW burnout was similar to the prepandemic rates. Nevertheless, HCW vaccination, engagement, motivation, teamwork, and loyalty subdimensions and their impact are still not well investigated during the pandemic.

#### 4.3.10 The HCW scientific development major dimension

Due to its importance, many studies aimed to evaluate HCW knowledge, attitudes, and practices (KAP) at the beginning of the pandemic [126]. HCW adherence to IC measures is affected by their KAP toward COVID-19 [127]. Some studies referred to insufficient knowledge about COVID-19 among nurses [128]. Surgeons were worried about losing their skills after months of lockdown due to paused practice [119]. However, HCWs were obliged to learn digital health skills and effectively communicate with patients during the pandemic [119].

A study [129] found that COVID-19 and non-COVID-19 publication productivity correlates with some factors. For example, epidemiologic, healthcare system-related, and pre-COVID publication expertise factors. Therefore, countries with a stable scientific infrastructure appear to maintain non-COVID-19 publication productivity nearly per year level. More incentives have to be drawn by HCO toward their HCW to encourage research and scientific productivity related to COVID-19.

#### 4.3.11 The technology and information system major dimension

Experts emphasized the role of technology and information (TI) in tackling COVID-19 as inevitable due to its importance in the response, prevention, preparedness, and recovery phases [130, 131]. TI system application varies from allowing HCO to maintain and share studies to producing different reports and follow-up with pandemic analysis. Telehealth is another example that proved helpful in the pandemic. It allowed HCWs to provide care for patients without direct physical contact, especially to patients at quarantine, while keeping them safe [132].

Researchers summarized the emerging technologies used to mitigate the threats of COVID- 19 in the following categories: artificial intelligence/deep learning, big data analytics, high-performance computing (HPC) infrastructures, robots, 3D printing technology, digital contact tracing technology, blockchain [113], bioinformatics systems, telemedicine, mobile phone, decision support system, the IC system in HCO, online interactive dashboard/geographic information system (GIS), Internet of Things (IoT), virtual reality (VR), surveillance systems, and internet search queries [130, 131]

Governments, healthcare systems, and HCO need to keep updated with the emerging technologies in this field, allocate resources to invest in them, and develop the required skills in HCWs to utilize them properly.

#### 4.3.12 Community care and reputation (the external) major dimension

Social sustainability indicators (SSIs) for healthcare facilities facing a crisis can be ambiguous to define and apply, so SSIs have been organized under the broad categorical concerns of well-being, values, agency, and inequality [133]. Despite the doctor–patient confidentiality clause and the protection law for patient data privacy, the Department of Health and Social Care for England has relaxed the rules on sharing confidential patient data. It required HCO and the NHS to exchange patient information to help fight COVID-19 [134]. Moreover, COVID-19 patient data led to society breaching patient privacy in some countries [119, 135], which may have stigmatized those patients [119].

As mentioned earlier, a study [102] emphasized the importance of sustainable environmental practices for better HW management. The political situation was also considered an external influence during the pandemic. It was highlighted in a study [136] in the Palestinian territories, which referred to the COVID-19 situation in the presence of the Israeli military occupation to have a double epidemic effect, which eventually impacted the performance of the Palestinian health system HCO during the pandemic.

However, the community role, such as exemptions offered by HCO for poor patients and social responsibility and patient privacy concerns and HCO market shares in COVID-19, are still poorly investigated.

#### 4.3.13 The managerial tasks and assessment major dimension

Standard policies, procedures, the availability of written standardized guidelines, and delivery in full and on time were considered essential in tackling COVID-19 [6]. A lack of standardization capability and conflicting or irrational managerial decisions were deemed dissatisfactory factors for HCWs in the pandemic [119].

Planning and preparedness are also crucial managerial tasks. The CDC developed a checklist to help hospitals assess and improve their preparedness for responding to COVID-19 [137]. Hospitals utilized a collection of some of the previously explained KPIs and dimensions to perform planning and internal assessment of their performance [133, 138].

Few studies [139, 140] have examined centralized governance’s impact on HCO during the pandemic, which positively affected reactive strategies. Learning from past pandemics also positively influences proactive and reactive strategies. However, the role of PE internally, such as using BSC or MBNQA tools, or external assessments, such as Joint Commission International (JCI) accreditations, ISO certification, auditing, or peer review on HCO during the pandemic, still requires more investigation.

### 4.4 Strengths and weaknesses

We believe that this paper has several strengths. First, this systematic review includes all types of studies with BSC implementations, such as books, theses, conference papers, letters to the editor, etc. Second, this review contains all implementations despite the country, language, or HCO administrative type, which gives an advantage of generalizing HCO results worldwide. Third, unlike other BSC reviews [35, 37], which included definitions of biobanks, pharmacies, laboratories, radiology, and medical colleges in HCO, this review limited the report to primary, secondary, or tertiary healthcare organizations. However, an initial assessment by top management to evaluate the importance of each dimension and KPI based on the health organizations’ strategy could be needed. This strategy leads to the homogeneity of the resulting studies and leads to more valid comparisons among the results. Fourth, this review calculates the use frequency of perspectives and the weights/importance assigned to them. Fifth, the first review has uniform the KPIs in homogenous major dimensions and subdimensions despite the categorization differences among implementations, yielding more precise results. Finally, this study is the first to analyze the implications of BSCs in HCO during the pandemic. The resulting KPIs and dimensions at this review can be generalized or replicable to other HCOs and hospitals.

On the other hand, this systematic review has some limitations. First, unlike previous studies, it excludes some HCO, such as laboratories, pharmacies, radiology departments, and biobanks, as specified in the inclusion/exclusion criteria. However, this will allow better concentration on KPIs directly related to HCO offering primary, secondary, and tertiary medical services. It is recommended for future reviews to study the other excluded types separately. Second, it includes only articles that report the complete implementation of BSC while excluding studies that display only the BSC design without reporting the full implementation results. Third, we extracted the KPIs from all resulting implementations despite their RoB. Future systematic reviews are encouraged to reperform when the number of studies with low and medium RoB is higher.

## 5. Conclusion

In conclusion, our review shows that the most frequently used perspectives in BSC papers were internal, financial, patient, learning and growth, HCW, managerial, community, and stakeholder perspectives. The perspectives that had the highest importance were internal, financial, learning and growth, patient, HCW, community, managerial, and stakeholder.

Moreover, this review solves the dilemma of the KPI categorization difference between BSC implementations by dimension unification into 13 major dimensions. The financial, efficiency and effectiveness, availability and the quality of supplies and services, managerial tasks, HCW scientific development, error-free and safety, time, HCW-centeredness, patient-centeredness, technology and information system, community care and reputation, HCO building, and communication. The proper utilization of the 13 major dimensions and the 45 subdimensions will serve as a planning, monitoring, evaluation, and continuous improvement tool for HCO, resulting in performance augmentation.

This research showed a lack of BSC utilization and any holistic PE approach in HCO during the COVID-19 pandemic. Additionally, some dimensions that are essential for PE are still poorly investigated. Future research for comprehensive PE of HCO during COVID-19 is required, which will lead to performance enhancement and mitigate the consequences of the pandemic on HCO.

## 6. Abbreviations

(AACI): American Accreditation Commission International
(AFT): Accelerated Failure Time
(ARDS): Acute Respiratory Distress Syndrome
(BSC): Balanced Scorecard
(CDC): Centers for Disease Control and Prevention
(COVID-19): Coronavirus Disease 2019
(CSR): Community Social Responsibility
(CSSD): Central Sterile Supply Department
(DEA): Data Envelopment Analysis
(DRG): Diagnostic Related Groups
(EFQM): European Foundation for Quality Management
(ER): Emergency Room
(GIS): Geographic Information System
(HAIs): Health-Associated Infections
(HCO): Health Care Organizations
(HCQI): Health Care Quality Indicator
(HCW): Health Care Workers
HMIS: Health Management Information System
(HPC): High-Performance Computing
(HW): Health Waste
(IC): Infection Control
(ICU): Intensive Care Unit
(IoT): Internet of Things
(ISO): International Organization for Standardization
(JCI): Joint Commission International
(KAP): Knowledge, Attitude, and Practices
(KPIs): Key Performance Indicators
(LOS): Length Of Stay
(MBNQA): Malcolm Baldrige National excellence model
(MBO): Management by Objectives
(MCDA): multiple-criteria decision analysis
(MeSH): Medical Subject Headings
(MMA): multimechanism approach
(MS): multistate
(NHS): National Health System
(OECD): Organization for Economic Co-operation and Development
(PATH): Performance Assessment Tool for quality improvement in hospital
(PE.): Performance Evaluation
(PPC): Physician–Patient Communication
(PPE): personal protective equipment
(PRISMA): Preferred Reporting Items for Systematic Reviews and Meta-Analyses
(QOPI): Quality Oncology Practice Initiative
(ROA): Return on Assets
(RoB): The Risk of Bias
(ROI): Return On Investment
(SAQ): Safety Attitude Questionnaire
(SQA): Singapore Quality Award
(SSI): Social Sustainability Indicators
(TC): Truncation Corrected
(T.I.): Technology and information
(TOPSIS): Technique for Order of Preference by Similarity to Ideal Solution
(TQIP): Trauma Quality Improvement Program
(TQM): Total Quality Management
(UK): United Kingdom
(USA): United States of America
(VR): Virtual Reality
(WHO): World Health Organization
(WT): Waiting Time

## 7. Declarations

### Ethics approval and consent to participate

Not Applicable since it is a systematic review.

### Consent for publication

Not Applicable.

### Availability of data and materials

All data generated or analyzed during this study are included in this published article [and its supplementary information files]

### Authors contributions

This systematic review was planned and designed by F. A, S. L, S.H., I. B & D.E. Specifically, F. A & S. performed the processes of searching for eligible studies and data extraction. F. A & S. H performed the interpretation of the data. F. A completed the writing of the current systematic review, and all of the authors participated in revising the manuscript. S.L., I. B & D. E supervised this systematic review. Ultimately, all of the authors approved the final manuscript and agreed to be personally accountable for the author’s contributions and to ensure that the accuracy and integrity of any part of the work were appropriately investigated and resolved.

## Data Availability

The data underlying this article are available in the Appendices upon the request of the journal.

## Acknowledgments

We want to thank Dr. Vahideh Moghaddam, Diêgo Andrade, and Wang Zhe for their help in translating and data extraction of Persian, Spanish, and Chinese articles, respectively. Additionally, we would like to thank Dr. Duha Shellah for contributing to revising the manuscript.

## 7. Appendices

S1 Appendix: PRISMA checklist.

S2 Appendix: Search strategies in PubMed, Embase, Cochrane, Google Scholar.

S3 Appendix: Quality Assessment.

