## Supplementary material for "A Systematic Review: The Dimensions utilized in the Performance Evaluation of Healthcare- An Implication during the COVID-19 Pandemic": S3 Appendix

Risk of Bias using ROBINS-I for Non-Randomized Studies of Interventions (NRSI) studies (Quasi- experimental and obdervational):

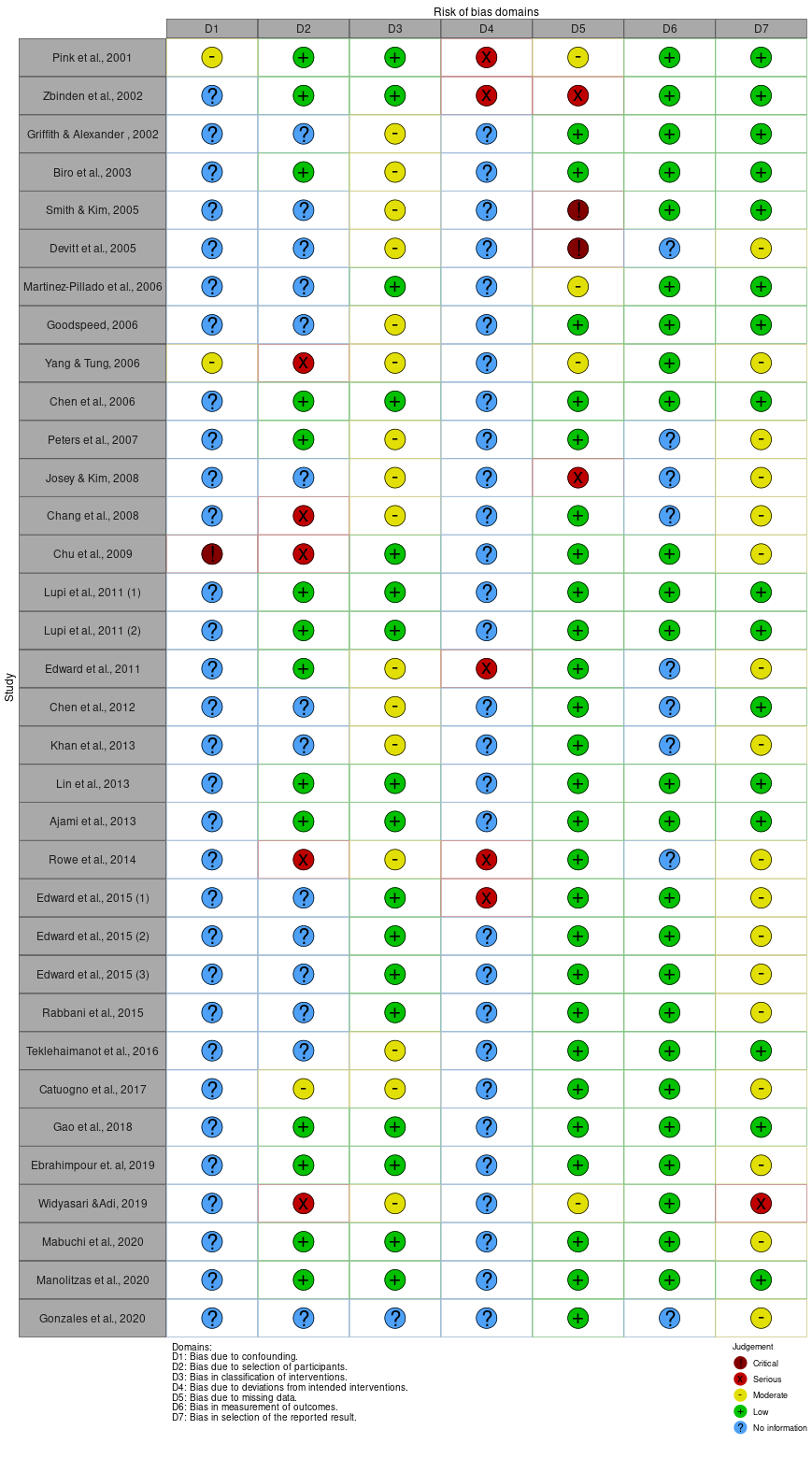

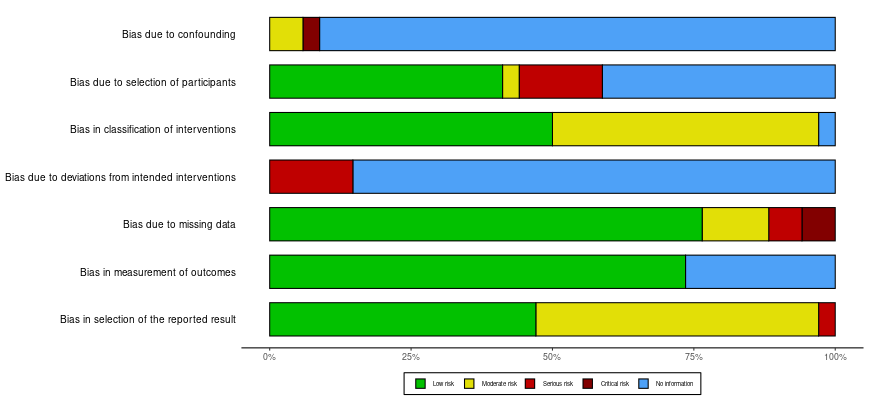

Risk of bias using ROB 2 for Randomized Controlled Trials (RCT) studies:

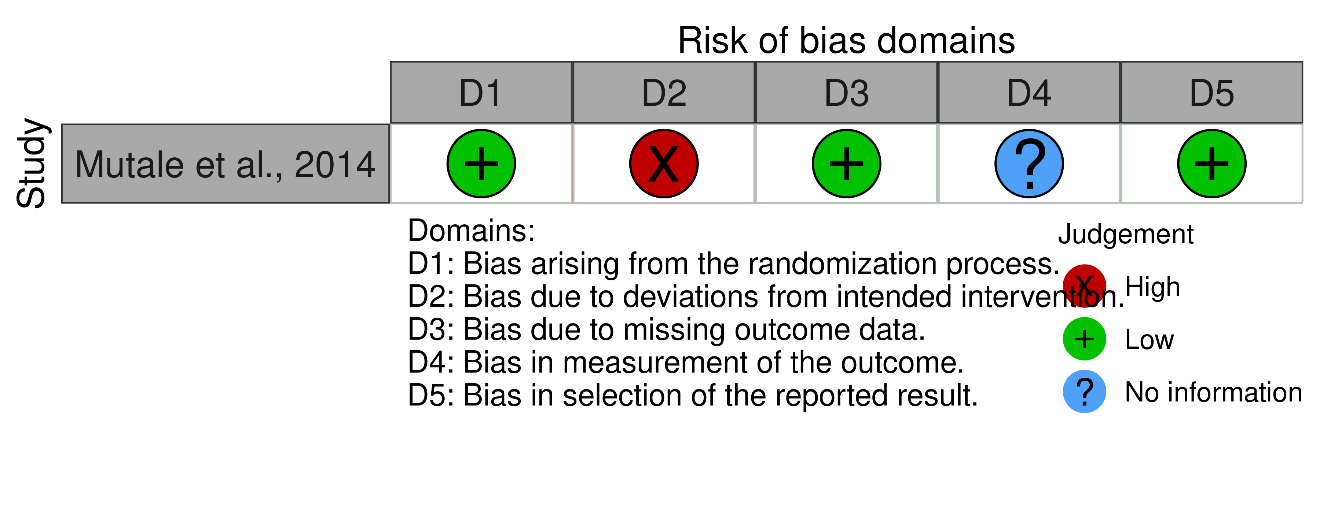
